# The Gut Metabolism is altered in patients with *CRB1*-associated inherited retinal degeneration

**DOI:** 10.1101/2024.02.22.24303210

**Authors:** Lude Moekotte, Joke H. de Boer, Sanne Hiddingh, Bram Gerritsen, Jutta Lintelmann, Alexander Cecil, L. Ingeborgh van den Born, Xuan-Thanh-An Nguyen, Camiel J.F. Boon, Maria M. van Genderen, Jonas J.W. Kuiper

## Abstract

**Purpose:** To compare the plasma metabolic profile of patients with a *CRB1*-associated inherited retinal degeneration (*CRB1*-IRD) with healthy controls (HCs).

**Design:** A case-control study.

**Methods:** Plasma concentration of 619 metabolites was measured with the MxP^®^ Quant 500 Kit in 30 patients with a *CRB1*-IRD and 29 HCs. We fitted a linear regression model with adjustments for age and sex based on the concentration of metabolites in µM (µmol/L), or on the sums and ratios of metabolites, to determine differences between patients and controls.

**Results:** Over-representation of pathways among metabolites associated strongest to *CRB1*-IRDs (*P* < 0.05, n = 62) identified amino acid pathways (such as beta-alanine, histidine, and glycine/serine) and bile acid biosynthesis, driven by a decrease in deoxycholic acid derivatives produced by gut microbiota. Enrichment analysis of metabolic classes across the plasma metabolic profile further identified significant positive enrichment for lipid metabolites glycerophospholipids, cholesterol esters, and ceramides, and significant depletion for bile acid metabolites. Further investigation of the sums and ratios (i.e., metabolism indicators) ascertained a significant decrease in intestinal microbial-dependent secondary bile acid classes.

**Conclusions:** Lipid metabolic alterations and decreased microbiota-related secondary bile acid concentrations indicate significant alterations in gut metabolism in patients with a *CRB1*-IRD.

## Introduction

Inherited retinal degenerations (IRDs) comprise a clinically and genetically heterogeneous group of monogenic retinal disorders, characterized by progressive loss of photoreceptors leading to severe visual impairment. Pathogenic variants in the *Crumbs homolog 1 (CRB1)* gene are associated with different IRD phenotypes, such as Leber congenital amaurosis, Severe Early Childhood Onset Retinal Dystrophy (SECORD), and cone-rod dystrophies.^1^ *CRB1* is highly expressed in retinal photoreceptors, as well as cells of the ciliary body that are involved in maintaining the blood-aqueous barrier.^2^ As compared to other retinal degenerations, *CRB1*-IRDs are more frequently associated with ocular inflammation or uveitis, suggesting that the disease may progress through inflammatory responses, and with loss of immune regulatory functions in the eye.^3–7^ This is supported by molecular studies, that show pro-inflammatory changes in peripheral blood T cell and dendritic cell populations, and elevated plasma complement components in peripheral blood.^8,9^ Interestingly, blood microbiota-produced metabolites detectable in blood, such as secondary bile acids, are associated with the severity of autoimmune uveitis.^10^ Retinal degeneration models also showed gut dysbiosis, which correlated with visual decline.^11,12^ The altered plasma metabolome might reflect differences in the gut microbiome, and can be used to probe disease mechanisms that involve gut metabolism and retinal function.

The retina is one of the most metabolic active areas in the body,^13,14^ of which photoreceptors have the highest energy demand.^15,16^ In retinal degenerations, a disturbance in glucose metabolism and oxidative damage contribute to photoreceptor death. This means that elucidating metabolic pathways responsible for the degeneration of photoreceptors is a major topic of interest.^17–19^ Previous studies in animal models of retinal degeneration showed that a metabolic switch in retinal cells leads to the release of neurotoxic inflammatory factors that damage photoreceptors.^20^ Other observations in animal models include decreased fatty acids in the retina which are correlated with photoreceptor degeneration.^21^ Also, the metabolism of amino acids, carbohydrates, nucleotides, and lipids has been proposed to play an important role in retinitis pigmentosa, the most common form of retinal degeneration.^22–26^ Oral supplementation to correct these metabolic changes proved to have a neuroprotective effect on the photoreceptor cells in mouse models of retinal degeneration.^27^ Studies of human retinal degeneration patients showed abnormalities in copper metabolisms,^28^ and lipid and amino acid metabolisms,^29–31^ suggesting that metabolic pathways may play a role in *CRB1*-IRDs.

Metabolomics is the multidisciplinary field studying the composition of metabolites (i.e., small molecules involved in cell metabolism) and has provided significant insights into immunomodulation and identification of metabolic and functional pathways that modulate clinical phenotypes.^32–35^ Application of metabolomics to retinal diseases has identified dysregulated functional pathways in proliferative diabetic retinopathy, age-related macular degeneration, and proliferative vitreoretinopathy,^22,36,37^ including alterations in fatty acids, amino acids, and nucleosides.^38–49^ Combining the current knowledge of immune system changes in *CRB1*-IRDs with new metabolic findings in patients can aid in better understanding the pathophysiological mechanisms of *CRB1*-IRDs. The purpose of this study was to elucidate the plasma metabolic profile of patients with a *CRB1*-IRD compared to healthy controls.

## Methods

### Patients

This study was performed in compliance with the guidelines of the Declaration of Helsinki and has the approval of the local Institutional Review Board (University Medical Center Utrecht (UMCU)). In total, 30 patients with a *CRB1*-IRD were included in the UMCU. Diagnosis was based on ophthalmic examination, imaging, and full field electroretinography. The diagnosis was confirmed with next-generation sequencing or whole-exome sequencing, displaying 2 or more pathogenic variants in the *CRB1* gene (**Supplementary Table 1**). All patients with systemic inflammatory conditions at the time of inclusion and/or patients with systemic immunomodulatory treatment were excluded from participation. On the day of inclusion, slit lamp examination was performed by an experienced uveitis specialist for assessment of vitreous cells and vitreous haze, and visual acuity was measured.

Twenty nine age matched healthy controls (HCs) with no known ophthalmic history or inflammatory disease were included through the research blood bank of the UMCU (known as the “Mini Donor Dienst”).

### Metabolomics

Venous blood samples were collected in Lavender Top Tubes ((#362084, BD Biosciences, Franklin Lakes, USA) of all patients and HCs on the day of inclusion (**Supplementary Table 2**). First, the tubes were centrifuged at 400g for 10 minutes. Plasma was transferred to a new tube, which was centrifuged at 1500g for 10 minutes. Next, the centrifuged plasma was collected in Micronic 1.4 mL round bottom tubes (#MP32022, Micronic, Lelystad, the Netherlands) and stored at −80 °C until measurement. For metabolomics measurement, the samples were shipped on dry ice to the *Metabolomics and Proteomics Core* (MPC) of the Helmholtz Facility München (Germany). Samples were randomized over one 96-wells plate and analyzed with the MxP^®^ Quant 500 Kit (#21094.12, Biocrates life sciences ag, Innsbruck, Austria). This kit is based on liquid chromatography-electrospray ionization-tandem mass spectrometric (LC-MS/MS) and flow injection-electrospray ionization-tandem mass spectrometric (FIA) measurements. In total, 5 aliquots of a plasma sample of an *in-house* plasma pool were used for quality control.

### Statistical analyses

Principal component analysis was performed with the *prcomp* function with the *factoextra* package.^50^ Metabolite concentrations of all plasma samples were expressed in units of µM (µmol/L). Individual metabolites and MetaboINDICATOR™ data – metabolite sums and ratios (Biocrates life sciences ag) – were analyzed separately. In case more than half of the data points of a metabolite were zero or a metabolite had zero variance, the metabolite was removed from the dataset. A Box Cox transformation was used to better approximate Gaussian distributions, after which all metabolite values were standardized relative to healthy controls. Then, a linear regression model was fitted with the *lm* function on each metabolite to analyze differences between patients with a *CRB1*-IRD and HCs, after which volcano plots were made with the *ggplot2* package to visualize the results.^51^ Metabolite classes were tested through a metabolite class enrichment analysis with the *GSEA* function of the *clusterProfiler* package.^52^ The classes were categorized as by the MxP^®^ Quant 500 Kit (#21094.12, Biocrates life sciences ag, Innsbruck, Austria) (**Figure 1A** and **Supplementary Table 2**). A *P* value adjusted with Benjamini Hochberg (BH) of < 0.05 was considered statistically significant. Over-representation analysis was performed with the *MetaboAnalystR* package v4.0.0. Other figures were generated with the *pie* function and *ggplot2* package.

**Figure 1.**
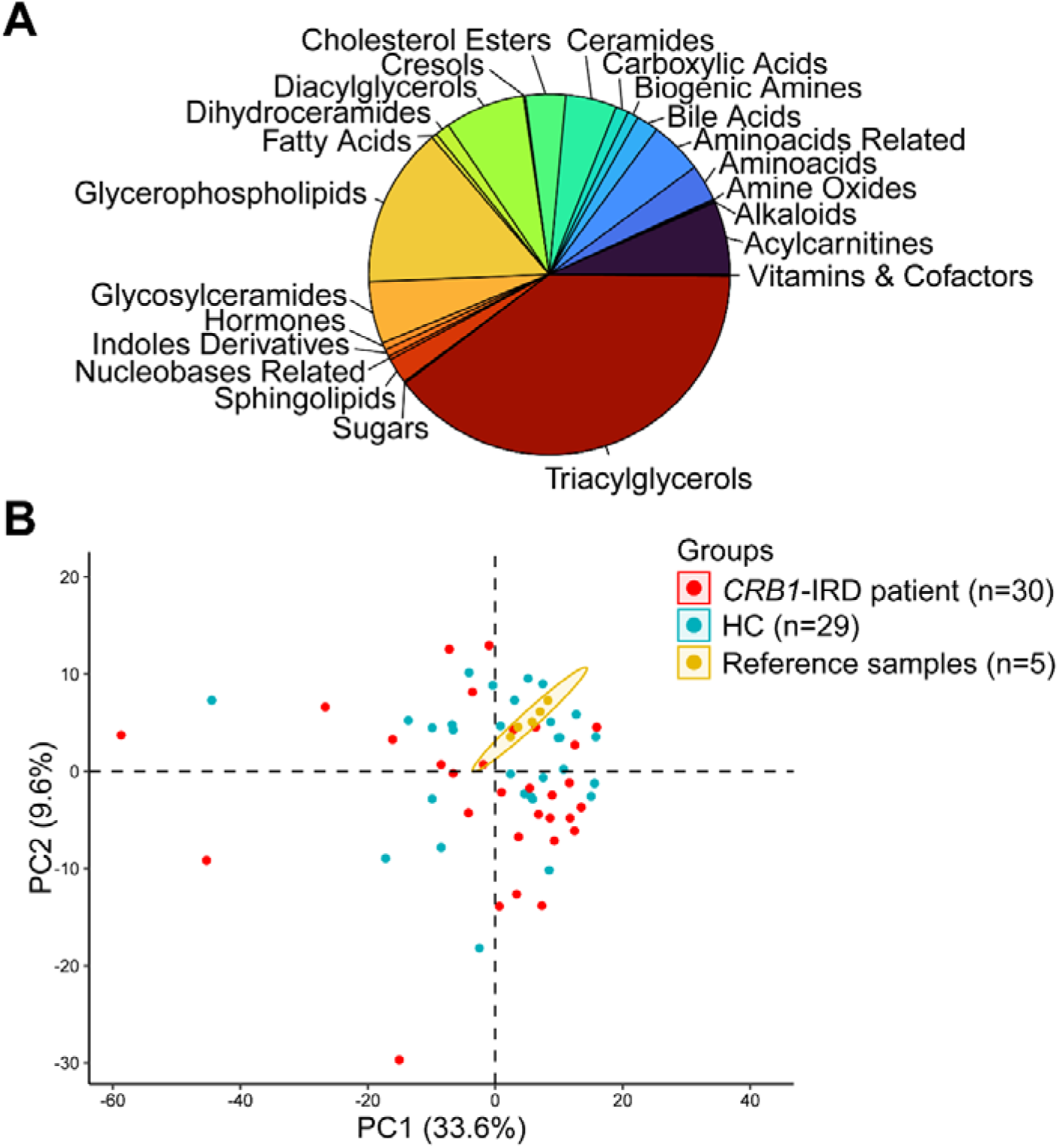
Overview and quality check. **A)** Pie chart representing coverage of the main classes of metabolites measured in plasma in this study. **B)** The first two principal components of metabolic profiles (n = 612 analytes; 7 analytes were removed in advance because more than half of the values were zero) in 30 patients with a *CRB1*-IRD and 29 healthy controls [HCs], and 5 (closely clustered) reference plasma samples.

## Results

We determined the concentrations of 619 metabolites across 23 metabolite classes in the peripheral blood plasma of 30 patients with a *CRB1*-IRD and 29 controls (**Figure 1A**). There were 7 metabolites in total that had more than half of their values zero, and they were removed. First, to visualize the variation of (reference) samples across the remaining 612 metabolites, we performed a principal component analysis, which showed close clustering of the reference plasma samples (**Figure 1B**).

Using a linear model, we compared patients with a *CRB1*-IRD and HCs, corrected for age and sex. We detected 62 lipids and small molecules that showed a nominal significant difference in concentration (**Figure 2A and C**). We found that patients had elevated levels of lipid metabolites, such as Cholesteryl Esters [CE(18:0), and CE(18:1)], sphingomyelins [SM C24:1] (**Figure 2A**), as well as amino acid derivatives Anserine and biogenic amine β-Alanine (**Figure 2C**). We further noted a decrease in plasma secondary bile acids (Glycodeoxycholic acid [GDCA], Deoxycholic acid [DCA], Taurodeoxycholic acid [TDCA], and Glycolithocholic acid [GLCA]) produced by the gut microbiota,^53^ cortisol (corticosteroid hormone), and nonessential amino acids, such as Alanine and Glycine (**Figure 2C**). Statistical differences at the level of individual metabolites did not exceed the threshold for statistical significance after correction for multiple testing (**Supplementary Table 3**). A statistical analysis of over-representation was performed to determine if metabolites associated with *CRB1*-IRDs were associated with specific metabolic pathways (49/62 nominal *P* < 0.05 present in libraries used in *MetaboAnalystR* analysis). This analysis revealed a significant enrichment for *Bile Acid Biosynthesis*, and amino acid metabolism pathways (i.e., *Beta-Alanine Metabolism*, *Glycine and Serine Metabolism*, *Histidine Metabolism*, and *Ammonia Recycling*) (**Figure 3A** and **Supplementary Table 4**).

**Figure 2.**
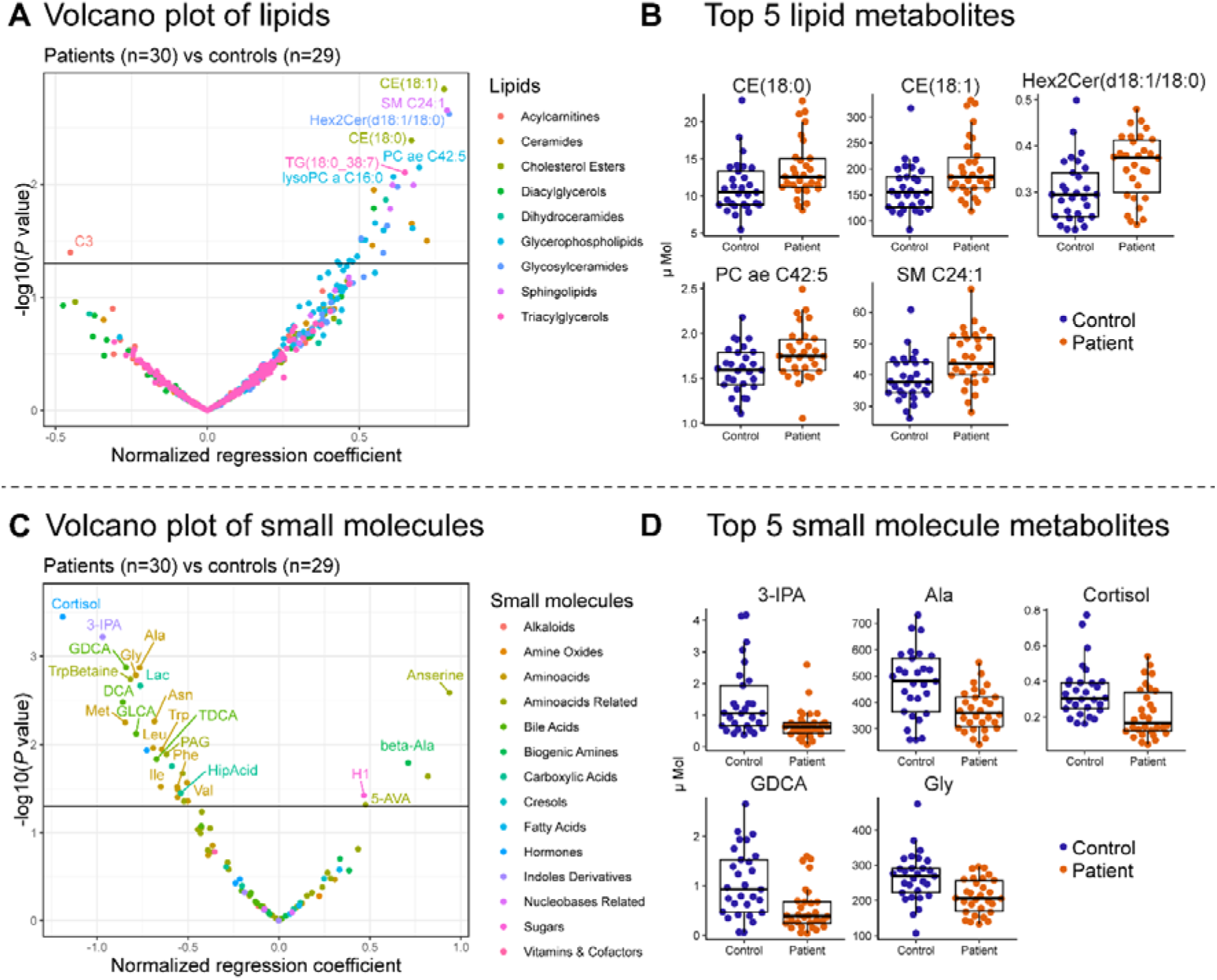
Changes in lipids and small molecules in plasma of patients with a *CRB1*-IRD. **A and C)** Volcano plot of the results from the comparison of patients versus controls. The normalized regression coefficient and the −log10 of the nominal *P* value are shown. The metabolites are colored per class of lipid as shown in the legend. The horizontal line represents a *P* value cutoff of 0.05. **B and D)** Boxplots of the top5 most significant metabolites (smallest *P* values). All top 5 lipid metabolites were increased in plasma of patients compared to HCs, while all top 5 small molecule metabolites were decreased.

**Figure 3.**
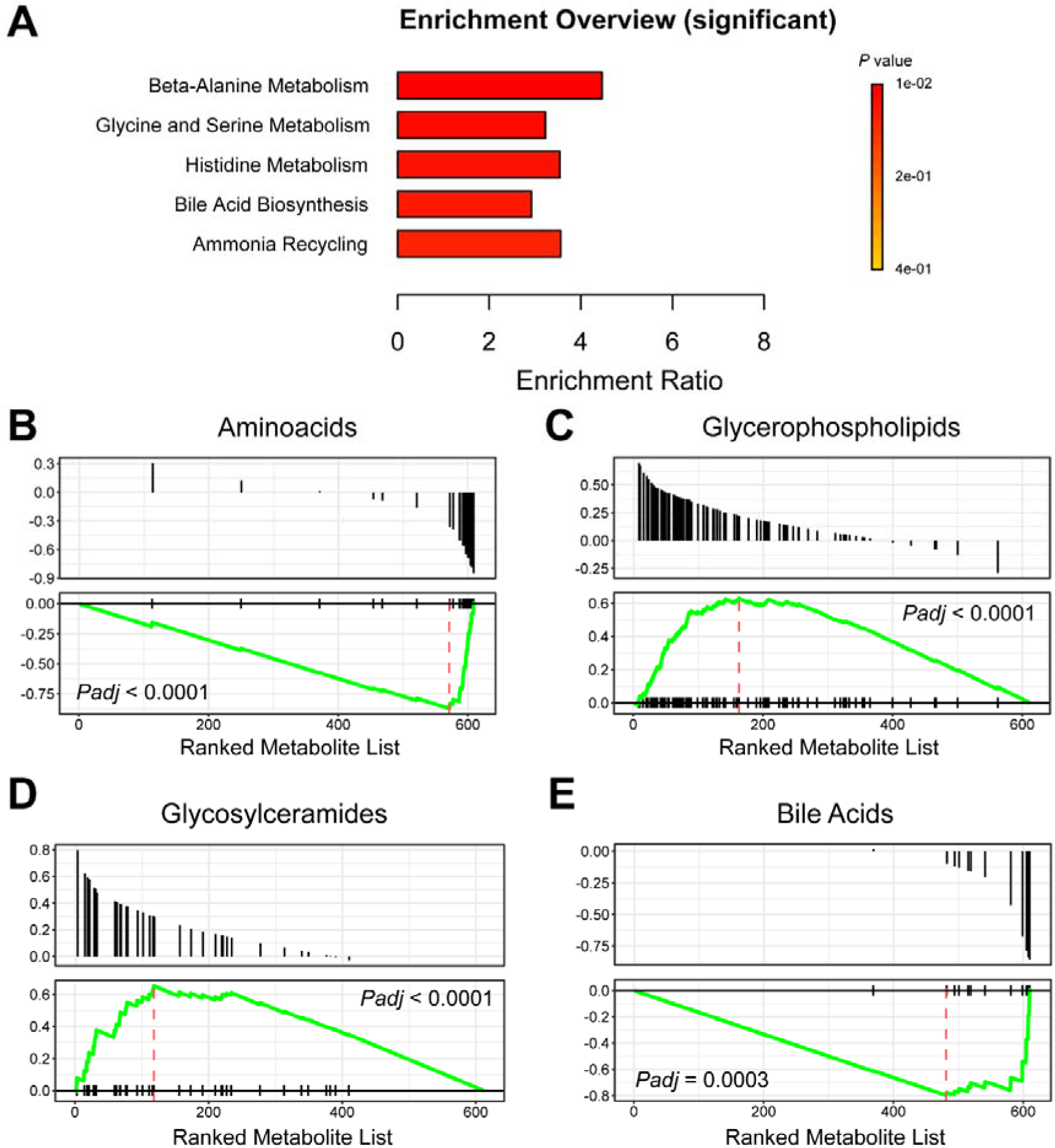
Enrichment and over-representation analysis. **A)** Over-representation analysis of 64 nominal significant metabolites. **B-E)** A metabolite class enrichment analysis performed on a ranked metabolite list. The top graph of each figure shows where the metabolite is located relative to the ranked list, as well as the distribution. The bottom graph of each figure visualizes the enrichment score. Graph B till E show the top 4 most significant metabolite classes. Glycerophospholipids (graph C) and Glycosylceramides (graph D) are positively enriched in patients, while the classes Aminoacids (graph B) and Bile Acids (graph E) are negatively enriched in patients.

**Figure 4.**
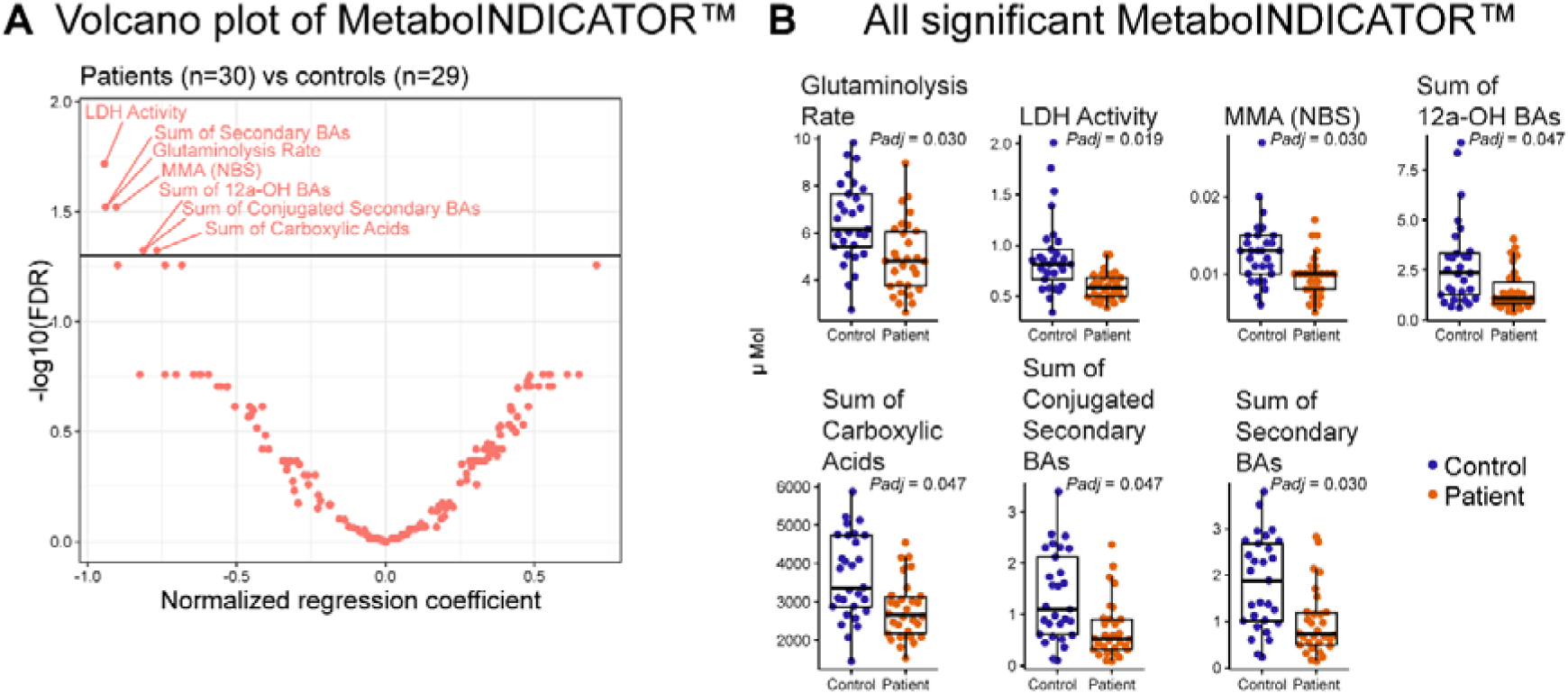
Linear model shows a significant difference in metabolic indicators associated with gut metabolism (MetaboINDICATOR™). **A)** Volcano plot visualizing the significance of metabolic indicators tested with a linear model. The normalized regression coefficient and the −log10 of the adjusted *P* value (FDR) is shown. The horizontal line represents a *Padj* value cutoff of 0.05. **B)** Boxplots of the significant metabolic indicators. All metabolic indicators are decreased in patients compared to HCs.

Next, we were interested to assess the enrichment for metabolic classes. Enrichment analysis considers the metabolic profile as a whole using differential data from every measured metabolite and searches for classes that display significantly coordinated changes in metabolite levels. To this end, we ranked the metabolic profiles (n = 612 metabolites) based on the statistics from the group comparison and performed metabolite class enrichment analysis for the 23 probed classes. After correcting for multiple testing, Bile Acids and Aminoacids were significantly and negatively enriched in the metabolome of patients (**Figure 3B and Figure 3E**). In addition, we detected a significant enrichment for Glycerophospholipids, Glycosylceramides, Sphingolipids, Cholesterol Esters, and Ceramides (**Figure 3B-E** for top 4 most significant and **Supplementary Table 5**).

Finally, we used the predefined sums and ratios of metabolites reported in clinical chemistry and biomedical phenotypes (MetaboINDICATOR™ data). Using a linear model corrected for sex and age, we detected a significant decrease in several metabolites produced by the intestinal microbiota, including Sum of Secondary Bile-Acids (*Padj* = 0.030),^54–57^ Sum of 12a-OH Bile-Acids (*Padj* = 0.047),^58–60^ Sum of Conjugated Secondary Bile-Acids (*Padj* = 0.047),^61,62^ and Sum of Carboxylic Acids (*Padj* = 0.047),^63^ as well as decreased Lactate dehydrogenase [LDH] Activity (*Padj* = 0.019) (**Supplementary Table 6 and 7** for description and references).^64,65^

## Discussion

In this study, we show differences in plasma metabolic profiles in patients with a *CRB1*-IRD compared to HCs. We detected enrichment for amino acid pathways, changes in lipid metabolites, and evidence for a decrease in secondary bile acids.

Bile acids are derivatives of cholesterol,^66–68^ and interest on bile acids in ocular diseases has peaked in the last decade. Bile acids have neuroprotective properties and protect against photoreceptor degeneration,^69–73^ mainly acting on oxidative stress and apoptosis.^70,74^ Primary and secondary bile acids influence metabolism and inflammation, and the gut microbiome is required to metabolize secondary bile acids from primary bile acids produced by the liver. Some intestinal species in the genus *Clostridium* metabolize the secondary bile acids we found to be reduced in patients with a *CRB1*-IRD, including deoxycholic acid (DCA) and their derivatives.^75^ DCA and derivatives of DCA play important roles in immune cells by activation of bile acid receptors and are known to regulate innate immunity and T cell activation.^76^ This is of interest because we have shown substantial activation of innate immune pathways and changes in T cell populations in patients with a *CRB1*-IRD.^8^ Perhaps, patients with a *CRB1*-IRD may exhibit changes in the gut microbiome, which are proxied by the changes in metabolites, such as secondary bile acids. Gut dysbiosis has also been linked to visual dysfunction in animal models of retinal degeneration.^11^ Interestingly, treatment with the closely related isomers of secondary bile acids protects against retinal degeneration in a widely used mouse model for retinal degeneration.^73,77,78^ *CRB1*-IRDs are characterized by inflammation and involve both innate and adaptive pathways, with intraocular inflammation (or uveitis) being an unusually frequent manifestation in this type of retinal degeneration.^8,9^ In models of non-infectious uveitis, microbiome changes are a key pathological trigger.^79^ This is important, because experimental autoimmune uveitis is characterized by low microbiota-related secondary bile acids in blood, and selective restoration of the gut bile acid pool attenuates the severity of uveitis.^10^ The anti-inflammatory mechanism of bile acids involves deoxycholic acid [DCA], which directly inhibits dendritic cell activation in uveitis.^10^ Dendritic cell alterations have also been reported in *CRB1*-IRDs, and are shown to reflect disease activity.^5,8^ Whether gut dysbiosis, DCA, and dendritic cell activation play a role in disease progression or if this relates to the inflammatory processes observed in selected patients with *CRB1*-IRDs exhibiting intraocular inflammation requires further investigation.

In the current cohort of patients with a *CRB1*-IRD, we also found evidence for changes in other lipid metabolites, which is in line with other studies on retinal degenerations.^80–83^ Blood metabolites, including lipids, enter the eye through two diffusion barriers: the outer limiting membrane and the outer blood-retina barrier (BRB).^83,84^ The RPE and the Bruch’s membrane mediate the metabolite exchange between photoreceptors and the blood, of which even large metabolites can cross and expand to all neural retinal layers.^83,85–87^ One notable finding in our study is the increase of cholesteryl esters in patients, which is the inactive form of cholesterol that transports and stores cholesterol.^88–92^ Cholesterol has toxic effects if not esterified,^91^ and higher concentrations of cholesteryl esters have been described in neurodegeneration after induced excitotoxic injury, Huntington’s disease, ALS (amyotrophic lateral sclerosis), MS (multiple sclerosis), and Alzheimer’s disease.^92–96^ When cholesterol excess occurs, esterification seems to occur to prevent further degeneration. In AMD patients, cholesteryl esters accumulate in the Bruch’s membrane and, together with unesterified cholesterol, represent >40% hard druse volume.^97^ It is unclear what the role of cholesteryl esters are in *CRB1*-IRDs, but with its protective effects it is possible that esterification of cholesterol occurs as a reaction to photoreceptor damage to limit further degeneration of photoreceptors.

Another significantly positively enriched lipid class in patients is Glycerophospholipids. In our metabolomics panel, all glycerophospholipids were lysophosphatidylcholines (LPCs). LPCs result from inflammatory processes and have been contributed to pro-inflammatory, as well as anti-inflammatory effects.^98,99^ They can increase the phagocytic activity of macrophages,^98^ increase genes involved in cholesterol biosynthesis,^100^ and increase the reactive oxygen production in polymorphonuclear leukocytes.^101^ A decrease of *Lpcat1* expression, coding for the protein lysophosphatidylcholine acyltransferase 1, is linked to photoreceptor degeneration in *rd11* mice.^102^ In diabetes type 1 and type 2 mice, *Lpcat1* is downregulated.^103^ Treating hyperlipidemic patients with an antioxidant significantly reduced LPCs in low-density lipoprotein compared to controls.^104^

In our study we included 30 *CRB1*-IRD patients and 29 HCs, limiting the power of our analyses. After correcting for multiple testing, the differences for individual metabolites did not exceed the statistical significance threshold. However, reflecting the physiological function of multiple metabolites functioning in a particular metabolic pathway, testing for grouped metabolic classes clearly showed significant enrichment after multiple testing correction for several pathways. The design of this cross-sectional study does, however, limit our ability to establish causality.

In conclusion, patients with a *CRB1*-IRD show an increase of lipid metabolites that are linked to photoreceptor degeneration, as well as a decrease of neuroprotective secondary bile acid concentrations, which indicate significant changes in the gut metabolism of patients with a *CRB1*-IRD. In the future, our findings combined with a larger prospective cohort study could aid us to elucidate causality and determine possible metabolic targets for therapy.

## Data Availability

All data produced in the present study are available upon reasonable request to the authors

## Acknowledgements

We would like to thank Dr. N.H. ten Dam-van Loon, Dr. J. Ossewaarde-van Norel, and Dr. V. Koopman-Kalinina Ayuso for their assistance performing slit lamp examination and assessment of vitreous cells and vitreous haze on all included patients.

